# Association between mild traumatic brain injury, brain structure, and mental health outcomes in the Adolescent Brain Cognitive Development Study

**DOI:** 10.1101/2022.06.02.22275940

**Authors:** Daniel A. Lopez, Zachary P. Christensen, John J. Foxe, Laura R. Ziemer, Paige R. Nicklas, Edward G. Freedman

## Abstract

**Background:** Children that experience a mild traumatic brain injury (mTBI) are at an increased risk of neural alterations that can deteriorate mental health. We test the hypothesis that mTBI is associated with behavioral and emotional problems and that structural brain metrics (e.g., volume, area) meaningfully mediate the relation in an adolescent population.

**Methods:** We analyzed behavioral and brain MRI data from 11,876 children who participated in the Adolescent Brain Cognitive Development (ABCD) Study. Mixed-effects models were used to examine the longitudinal association between mTBI and mental health outcomes. Bayesian methods were used to investigate brain regions that are intermediate between mTBI and symptoms of poor mental health.

**Results:** There were 199 children with mTBI and 527 with possible mTBI across the three ABCD Study visits. There was a 7% (IRR = 1.07, 95% CI: 1.01, 1.13) and 15% (IRR = 1.16, 95% CI: 1.05, 1.26) increased risk of emotional or behavioral problems in children that experienced possible mTBI or mTBI, respectively. Possible mTBI was associated with a 17% (IRR: 1.17, 95% CI: 0.99, 1.40) increased risk of experiencing distress following a psychotic-like experience. We did not find any brain regions that meaningfully mediated the relationship between mTBI and mental health outcomes. Analysis of volumetric measures found that 3 to 5% of the total effect of mTBI on mental health outcomes operated through total cortical volume. Image intensity measure analyses determined that 2 to 5% of the total effect was mediated through the left-hemisphere of the dorsolateral prefrontal cortex.

**Conclusion:** Results indicate an increased risk of emotional and behavioral problems in children that experienced possible mTBI or mTBI. Mediation analyses did not elucidate the mechanisms underlying the association between mTBI and mental health outcomes.

## 1. Introduction

In 2014, there were more than 812,000 cases of pediatric traumatic brain injury (TBI) that required an emergency department visit in the United States (1). The most common mechanisms of injury for pediatric TBI included falls, being struck by or against an object, and motor vehicle accidents (2). Mild TBI (mTBI) accounts for about 75% of all TBIs and typically involves an external force to the head that in turn alters brain function (2-4). Additional diagnostic criteria include loss of consciousness (LOC) and post-traumatic amnesia (5). Approximately 14% of children with mTBI experience disability that requires specialized medical and educational services (6). For most children, mTBI symptoms persist for 1 to 3 months after injury (7). A minority of children experience long-term consequences from mTBI that include psychiatric illness (e.g., psychosis and emotional problems) (8-10). Whether any structural brain metrics mediate the association between mTBI and psychiatric sequelae is unclear. Detection of structural abnormalities can improve our understanding of the mechanisms underlying the relationship.

Pathophysiological explanations for the psychological and emotional consequences of mTBI are not fully understood. Explanations focus on damage to brain regions typically associated with behavioral functioning (e.g., frontal and temporal lobes) (9). Findings from imaging studies include decreased volume in the left lateral frontal cortex and left inferior frontal gyrus of participants that developed major depression following TBI (11). Injury severity can also play a role in structural abnormalities. A study on the effects of TBI in children (mean age at injury = 6.59) found that more severe injuries led to a reduction in gray matter (GM) and white matter (WM) volumes, while less severe injuries were associated with increased cerebrospinal fluid volumes and reduced hippocampal volume suggesting that milder cases may be harder to visualize using magnetic resonance imaging (MRI) (12). Longitudinal findings support changes during the first year post-injury that include volume decreases in the anterior cingulate WM, left cingulate gyrus WM, and right precuneus GM (13).

Psychotic-like experiences (PLEs) are a range of hallucination and delusion symptoms that occur in the general population and can be used to detect preclinical risk of psychosis (14, 15). PLEs have been found to be associated with various structural changes in the brain, including reduced cortical volumes, increased amygdalar volume, and increased mean diffusivity in white matter (16). A case-control study conducted specifically with adolescents found a significant association between reported PLEs and changes in white matter microstructure (17). A separate case-control study reported similar abnormalities in white matter microstructure in young adults with psychotic experiences (18). A study with children aged 11 to 13 found that psychosis risk was associated with GM increases in some gyri, volume decreases in others, and WM decreases along multiple tracts (19). A cohort study in children aged 10 to 13 found associations between subclinical psychotic experiences and significant volumetric increases in the left hippocampus, right caudate, right lateral ventricle, and left pallidum (20). Importantly, another study using cross-sectional data from the Adolescent Brain Cognitive Development (ABCD) Study found neural changes that were associated with PLEs in young children (21). The wide range of findings of brain associations with psychosis risk and symptoms further demonstrates the need to understand other risk factors (e.g., history of TBI) that could be involved in the relationship.

Behavioral and emotional disturbances are an additional domain in the realm of post-TBI sequelae (22). Pathophysiological explanations for these disturbances include damage to brain regions involved in neurocognitive development (e.g., frontal lobes, temporal lobes, cingulate regions) (23, 24). The damage to neural substrates can manifest into persistent behavioral and emotional deficits (25). A cross-sectional study using a pediatric population aged 12 to 17 found an increased risk of depression in children that experienced a concussion (OR = 3.3, 95% CI: 2.0, 5.5) (26). Separate studies concluded that TBI was similarly associated with an increased risk of anxiety disorders (27), secondary ADHD and aggression (28), and other mental health difficulties (29). Longitudinal findings support a long-lasting effect of TBI on behavioral and emotional domains. A pediatric study in children aged 6 to 15 reported that 60% of their sample developed a novel psychiatric disorder following TBI (30). Approximately 75% of novel cases had persistent psychiatric disorders at least 1-year post-injury (30). Scales of global psychological impairment also point towards reduced emotional and behavioral functioning post-TBI. A pediatric study in the United States found that 37% of children with moderate or severe TBI had borderline/clinical elevations in social and behavioral problems (31). The study used the Child Behavior Checklist (CBCL) to assess total problems across syndrome scales representing multiple domains of mental health (31). Altogether, there is evidence that TBI and head injuries are implicated in poor mental health outcomes.

In the current study, we first examined the longitudinal association between mTBI and mental health outcomes in the ABCD Study. We hypothesized that a history of possible mTBI and mTBI would result in 1) an increased risk of experiencing distress due to a psychotic-like experience and 2) an increased risk of experiencing emotional or behavioral problems. We then assessed whether structural MRI (sMRI) measures mediated the association between mTBI and separate mental health outcomes. We hypothesized that structural brain metrics (e.g., total cortical volume) would meaningfully mediate the association between mTBI and mental health outcomes. The current study was conducted using data from the ABCD 4.0 data release.

## 2. Materials and Methods

### 2.1. Sample

The ABCD Study is a prospective longitudinal study that enrolled 11,876 children aged 9-10 at 21 research sites throughout the United States. The ABCD Study plans to conduct yearly in-person visits until participants are at least 19 to 20 years old (32). Exclusion criteria for the ABCD Study are discussed elsewhere (33). Notably, exclusion criteria for enrollment included a history of severe traumatic brain injury (i.e., TBI with LOC greater than 30 minutes), Amnesia greater than 24 hours, positive neuroimaging findings secondary to TBI, a current diagnosis of schizophrenia, or an MRI contraindication (e.g., claustrophobia).

ABCD Study data were accessed using the National Institute of Mental Health Data Archive. The measures in the current analysis include data collected at the baseline (n=11876), 1-year (n=11225), and 2-year (n=10414) follow-up visits. Results are reported for separate models using complete-case analysis and multiple imputation for missing data. Parent consent and child assent were approved by the central Institutional Review Board at the University of California – San Diego. Study procedures were conducted in accordance with the declaration of Helsinki. Code for the replication of study results can be obtained on Github: https://github.com/Daniel-Adan-Lopez/ABCD_mTBI/blob/main/Rcode

### 2.2. Prodromal Psychosis Scale

The ABCD Prodromal Questionnaire – Brief Child Version (PQ-BC) is a measure of psychotic-like experiences (PLEs) during childhood (33). The PQ-BC was adopted from the PQ-B measure created for adults (34). PLEs are considered a sub-threshold, non-clinical symptom of psychosis (15). The 21-item PQ-BC has previously been assessed as a valid self-report instrument for children and adolescents (33). Participants completed the PQ-BC at the annual study visit (i.e., at three time points in the current analyses). Participants were asked by an interviewer whether they had experienced a PLE (e.g., auditory hallucinations) in the past month. Endorsement of a PLE resulted in a follow-up question related to distress caused by the event. Children that experienced distress (i.e., answered yes) then were asked to describe the event using a 5-point Distress scale (1=Not very bothered, 2=Slightly bothered, 3=Moderately bothered, 4=Very much bothered, 5=Extremely bothered). Distress scores were calculated as the total number of endorsed questions weighted by level of distress (35). Distress scores ranged from 0 to 126 (33).

### 2.3. Child Behavior Checklist

The parent-reported Child Behavior Checklist (CBCL) is a broad measure of psychological functioning in children (36). The CBCL has previously been shown to capture five spectra of psychopathology in the ABCD Study: internalizing, somatoform, detachment, externalizing, and neurodevelopmental (37). Parents completed the 112-item CBCL at the annual study visit (i.e., at three time points in the current analyses). Parents answered individual items that describe the child on a 3-point Likert scale (0=Not True, 1=Somewhat/Sometimes True, 2=Very True/Often True). The analyses used the CBCL Total Problems raw score. The Total Problems score encompasses the sum of all responses on the syndrome scales of the CBCL (i.e., Anxious/Depressed, Withdrawn/Depressed, Somatic Complaints, Social Problems, Thought Problems, Attention Problems, Rule-Breaking Behavior, Aggressive Behavior) (36). CBCL Total Problems scores ranged from 0 to 224.

### 2.4. Traumatic Brain Injury

The Ohio State Traumatic Brain Injury Screen - Short Modified (OTBI) is a measure of lifetime exposure to TBI (38). The measure has previously been assessed for reliability and validity in longitudinal studies (38, 39). The baseline OTBI included four questions directed at the parent or guardian assessing past head injury and consequent LOC or other complication (e.g., feeling dazed or gap in memory from the injury). Follow-up ABCD visits included a revised OTBI that assessed injury since the last study visit. TBI was then determined according to the following OTBI summary indices: Improbable TBI (no TBI or TBI without LOC or memory loss); Possible mild TBI (TBI without LOC but memory loss); Mild TBI (TBI with LOC ≤ 30 minutes); Moderate TBI (TBI with LOC 30 minutes to 24 hours); Severe TBI (TBI with LOC ≥ 24 hours). Cases of moderate (n=9) and severe TBI (n=3) were removed to reduce the influence of small samples. The longitudinal TBI variable included levels for no TBI (reference), possible mild TBI, and mild TBI.

### 2.5. Image acquisition

Children enrolled in the ABCD Study underwent a sMRI scan at the baseline and 2-year visit (40). 3d T1-weighted and T2-weighted images were acquired with 3 tesla(T) scanner platforms (Siemens Prisma and Prisma Fit, GE MR 750, and Philips Achieva dStream and Ingenia) (41). Scanner type varied by ABCD Study site. Cortical surface reconstruction of MRI data was performed using FreeSurfer v5.3 (40). A modified FreeSurfer pipeline included quality control measures focused on accuracy of cortical surface reconstruction (40). An atlas-based, volumetric segmentation procedure was used to label subcortical structures for the identification of regions of interest (ROIs). The current analyses examined the indirect effect of structural brain metrics and image intensity measures on mental health outcomes. Structural brain metrics included cortical thickness, area, volume, and sulcal depth. Image intensity measures included T1w and T2w cortical contrast measures that were calculated using GM and WM values ([white-gray]/[white+gray]/2) (40). The gray-white contrast (GWC) is included as a potentially sensitive indicator of psychopathology (42). Quality control and processing of sMRI data were initially performed by the ABCD Data Analysis, Informatics and Resource Center and included trained technician review of the accuracy of cortical surface reconstruction (40).

### 2.6. Statistical analysis

The longitudinal data analyses included a mixed-effects model that accounted for 1) clustered data (i.e., within-individual correlation across repeated measurements), 2) a nested structure (e.g., family units nested within study sites), 3) a skewed outcome. All analyses using the imputed and non-imputed data were conducted using the R glmmTMB package (43, 44). Study visit was included as both a fixed and random effect (i.e., random intercept and slope for each study participant). The multilevel model included a random effect for the family unit nested within the ABCD study site. A negative binomial model was considered appropriate to account for the discrete count data of the PQ-BC Distress and CBCL-Total Problems scores. Models were compared using the Akaike’s Information Criterion. Covariates that did not improve the model (i.e., did not decrease the criterion value) were removed to avoid problems with overfitting (45).

The final model included adjustment for the following participant characteristics: age, sex, race/ethnicity, perceptions of neighborhood safety, perceptions of parental monitoring, and whether a sibling (e.g., twins) was enrolled in the study. Adjustment for family characteristics included household income, parental education, parent marital status, and family history of psychosis. Results are expressed as incidence rate ratios (i.e., the exponentiated beta coefficient) and 95% confidence intervals using a likelihood profile (44).

#### 2.6.1. Mediation Analysis

A Bayesian multilevel mediation analyses was conducted to examine whether certain sMRI measures were involved in the association between mTBI and mental health outcomes. Scanner type and motion score were included as covariates in addition to previously mentioned variables. The mediation analyses included only data from the year-2 visit. The year-1 data was excluded from the mediation analysis due to the lack of MRI data (i.e., children did not complete an MRI scan). Baseline data was excluded to reduce the influence of recall bias (e.g., problem recalling a head injury that potentially occurred a decade ago) and time since head injury. Consequently, the mediation analysis primarily describes the neural mechanisms involved in recent mTBI cases and mental health outcomes. The multilevel analyses included a random intercept for study site and group-clustering by family (46). Brain measures and interview age were normalized before analysis. The indirect effect and the proportion mediated (i.e., indirect effect divided by total effect) along with the corresponding 95% credible intervals are reported for separate ROIs and GWCs. Path coefficients are reported for each mediation model. The multilevel mediation analyses were completed using the brms R package for the Bayesian regression models and BayestestR for summary of the mediation models (46, 47).

#### 2.6.2. Covariate assessment

The child and their parent or guardian completed a series of surveys at each study visit. Parent or guardian reported measures included child’s biological sex, child’s race/ethnicity, household income, household educational attainment, parental marital status, and family history of psychosis. The child was asked about their perceived neighborhood safety and parental monitoring. Perceived neighborhood safety was measured using the Neighborhood Safety/Crime Survey modified form. Children responded on a 5-point Likert scale whether they agreed with the statement “My neighborhood is safe from crime” (1= Strongly Disagree, 2 = Disagree, 3= Neutral, 4 = Agree, 5 = Strongly Agree). Parental monitoring was measured using the Parental Monitoring and Supervision form. The five-item measure asked children about the level of supervision of their parent or guardian using a 5-point Likert scale. Prior research has found that permissive parenting can predict worse behavioral outcomes post-TBI (23, 48). The analyses incorporated longitudinal change in repeated measures (e.g., a decrease in household income after baseline).

#### 2.6.3. Missing Data

The amount of missing household income data at each study visit was 8.6% at baseline, 8.2% at the year-1 visit, and 8.0% at the year-2 visit. Multilevel multiple imputation of missing data was conducted using the R mice package (49). First, the missing data mechanisms were examined to determine whether imputation was necessary. Second, 20 imputed data sets with 20 iterations and k=3 donors were created using a model that included all variables in the analysis model. The model included study site, the family unit, and visit number as both a fixed and random effect (i.e., a level 2 variable). Predictive mean matching was used to impute the missing household income values. Predictive mean matching is considered an accurate method of imputing ordered categorical variables (e.g., household income) in multilevel models (50). Finally, a pooled analysis was conducted using Rubin’s rule to estimate the parameters (51, 52).

## 3. Results

### 3.1. Sample description

The demographics of participants at the study visits are shown in Table 1. There were 199 cases of mild TBI and 527 cases of possible mild TBI reported across the three visits. A total of 30,111 observations were included in the PQ-BC model (10.2% of observations were excluded due to missing data). There were 30,113 observations included in the CBCL Total Problems model (10.2% of observations excluded due to missing data).

**Table 1.** Participant Demographics at ABCD Study Visits

|  | **Baseline**<br>**(n=11,876)** | **Year 1**<br>**(n=11,225)** | **Year 2**<br>**(n=10,414)** |
| --- | --- | --- | --- |
| **Traumatic Brain Injury** |  |  |  |
| No | 11415 (96.2) | 11072 (98.8) | 10217 (98.6) |
| Possible mild TBI | 322 (2.7) | 98 (0.9) | 107 (1.0) |
| Mild TBI | 128 (1.1) | 37 (0.3) | 34 (0.3) |
| **Child gender** |  |  |  |
| Female | 5680 (47.8) | 5353 (47.7) | 4962 (47.6) |
| Male | 6196 (52.2) | 5872 (52.3) | 5452 (52.4) |
| **Age of child (months (SD))** | 118.98 (7.50) | 131.09 (7.72) | 144.04 (7.95) |
| **Race/ethnicity of child** |  |  |  |
| Asian | 252 (2.1) | 241 (2.1) | 216 (2.1) |
| Black | 1784 (15.0) | 1597 (14.2) | 1422 (13.7) |
| Hispanic | 2411 (20.3) | 2222 (19.8) | 2086 (20.0) |
| White | 6180 (52.0) | 5989 (53.4) | 5602 (53.8) |
| Other | 1247 (10.5) | 1174 (10.5) | 1088 (10.4) |
| **Household Income (\$)** | | | |
| <50,000 | 3223 (29.7) | 2933 (28.5) | 2687 (28.0) |
| 50,000 to < 100,000 | 3071 (28.3) | 2941 (28.5) | 2769 (28.9) |
| ≥ 100,000 | 4564 (42.0) | 4434 (43.0) | 4129 (43.1) |
| Missing | 1018 (8.6) | 917 (8.2) | 829 (8.0) |
| **Household Educational Attainment** |  |  |  |
| < HS Diploma | 593 (5.0) | 526 (4.7) | 463 (4.5) |
| HS Diploma/GED | 1132 (9.5) | 1009 (9.0) | 910 (8.7) |
| Some College | 3079 (26.0) | 2849 (25.4) | 2620 (25.2) |
| Bachelor | 3015 (25.4) | 2896 (25.8) | 2742 (26.4) |
| Post Graduate Degree | 4043 (34.1) | 3933 (35.1) | 3667 (35.3) |
| **Parent Marital Status** |  |  |  |
| Married | 7990 (67.8) | 7676(68.9) | 7185 (69.4) |
| Not Married | 3790 (32.3) | 3472(31.1) | 3163 (30.6) |
| **PQ-BC Distress Score (mean (SE))** | 6.32 (0.10) | 4.62 (0.09) | 3.56 (0.08) |
| **CBCL Total Problems Score (mean (SE))** | 18.18 (0.17) | 17.54 (0.17) | 16.43 (0.17) |

Note: Distributions are represented as n (%) unless otherwise noted.
SD = Standard Deviation
SE = Standard Error

### 3.2. Prodromal Psychosis – Distress Score

The mean PQ-BC Distress score was 6.32 (SE = 0.10) at baseline, 4.62 (SD = 0.09) at year 1, and 3.56 (SD = 0.08) at year 2. The maximum observed score was 117. Greater PQ-BC Distress scores were evident in the following groups across the three time points (Table 2): 1) children from lower income households, 2) Black and Hispanic children, 3) children that reported low neighborhood safety, 4) children with a family history of psychosis, 5) children with an unmarried parent. PQ-BC Distress scores were not meaningfully different in male and female children (mean = 4.90 and 4.88, respectively).

**Table 2.**
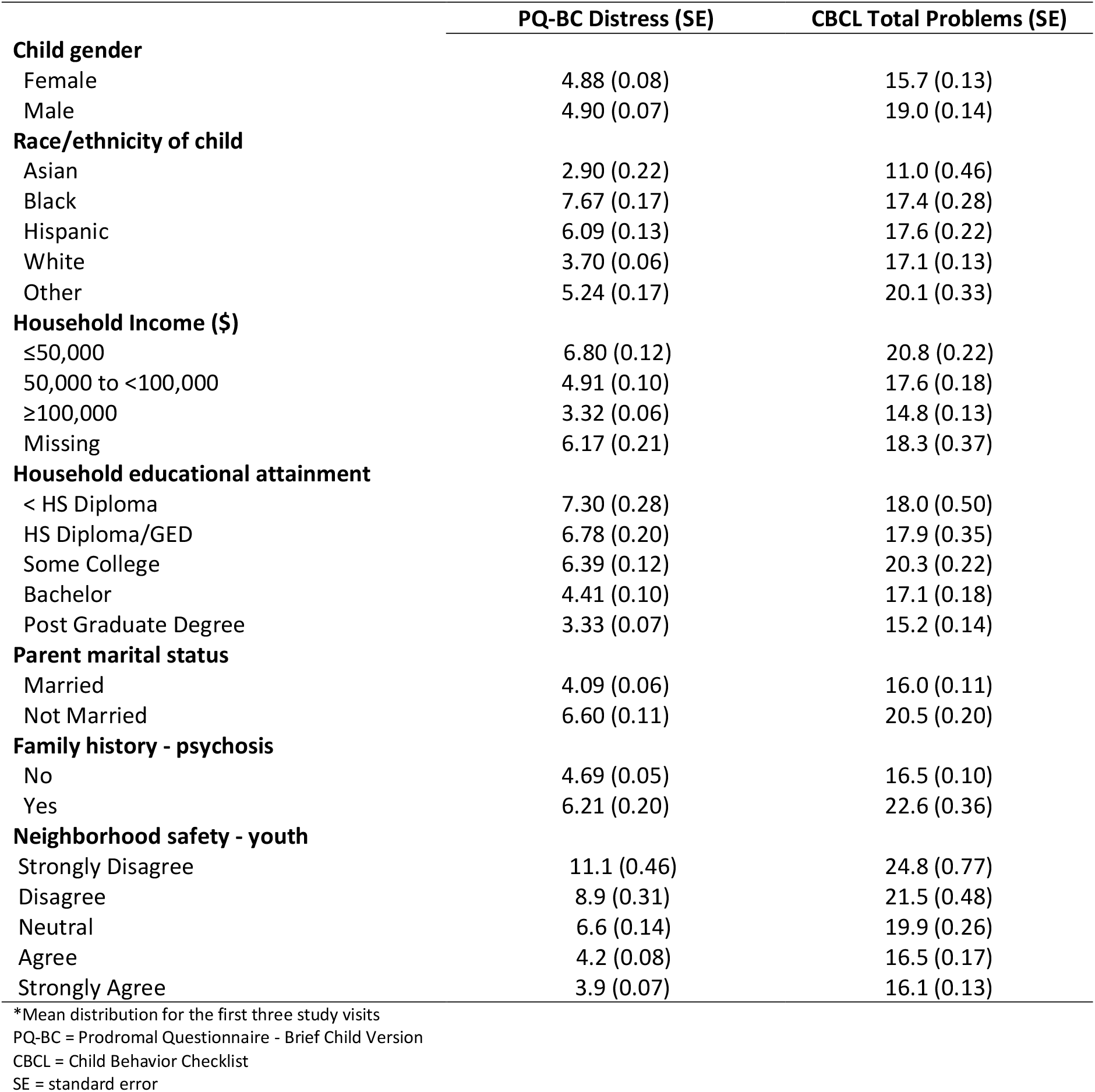
Mean PQ-BC and CBCL scores across participant characteristics*

### 3.3. Child Behavior Checklist – Total Problems Score

The mean CBCL Total Problems score was 18.18 (SE = 0.17) at baseline, 17.54 (SE = 0.17) at year 1, and 16.43 (SE = 0.17) at year 2. The maximum observed score was 161. Greater CBCL Total Problems scores were evident in the following groups across the three time points (Table 2): 1) male children, 2) children from lower income households, 3) children that reported low neighborhood safety, 4) children with a family history of psychosis, 5) children with an unmarried parent.

### 3.4. Traumatic Brain Injury

Males and children with a family history of psychosis were more likely to report experiencing a mTBI at the three study visits (Table 3). There was no significant difference in age and parental marital status between the two groups.

**Table 3.** Characteristics of Participants with a Mild TBI at ABCD Study Visits

|  | Mild Traumatic Brain Injury |  |  |  |  |  |
| --- | --- | --- | --- | --- | --- | --- |
|  | visit 1 |  | visit 2 |  | visit 3 |  |
|  | Yes (n=128) | No (n=11415) | Yes (n=37) | No (n=11072) | Yes (n=34) | No (n=10217) |
| **Sex** |  |  |  |  |  |  |
| Female | 52 (40.6) | 5500 (48.2) | 15 (40.5) | 5289 (47.8) | 13 (38.2) | 4885 (47.8) |
| Male | 76 (59.4) | 5915 (51.8) | 22 (59.5) | 5783 (52.2) | 21 (61.8) | 5332 (52.2) |
| **Age (months (SD))** | 119 (7.90) | 119 (7.49) | 132 (7.52) | 131 (7.71) | 146 (8.53) | 144 (7.95) |
| **Race/ethnicity (%)** |  |  |  |  |  |  |
| Asian | 0 (0.0) | 248 (2.2) | 0 (0.0) | 237 (2.1) | 0 (0.0) | 214 (2.1) |
| Black | 8 (6.2) | 1742 (15.3) | 3 (8.1) | 1581 (14.3) | 3 (8.8) | 1407 (13.8) |
| Hispanic | 30 (23.4) | 2337 (20.5) | 5 (13.5) | 2206 (19.9) | 11 (32.4) | 2049 (20.1) |
| White | 77 (60.2) | 5891 (51.6) | 21 (56.8) | 5896 (53.3) | 15 (44.1) | 5487 (53.7) |
| Other | 13 (10.2) | 1195 (10.5) | 8 (21.6) | 1150 (10.4) | 5 (14.7) | 1060 (10.4) |
| **Household Income (\$)** | | | | | | |
| ≤50,000 | 30 (23.4) | 3133 (27.4) | 7 (18.9) | 2903 (26.2) | 8 (23.5) | 2648 (25.9) |
| 50,000 to <100,000 | 31 (24.2) | 2950 (25.8) | 15 (40.5) | 2899 (26.2) | 10 (29.4) | 2709 (26.5) |
| ≥ 100,000 | 59 (46.1) | 4352 (38.1) | 13 (35.1) | 4362 (39.4) | 13 (38.2) | 4042 (39.6) |
| Missing | 8 (6.2) | 980 (8.6) | 2 (5.4) | 908 (8.2) | 3 (8.8) | 818 (8.0) |
| **Household educational attainment** |  |  |  |  |  |  |
| < HS Diploma | 5 (3.9) | 577 (5.1) | 2 (5.4) | 520 (4.7) | 0 (0.0) | 459 (4.5) |
| HS Diploma/GED | 7 (5.5) | 1108 (9.7) | 2 (5.4) | 1006 (9.1) | 2 (5.9) | 905 (8.9) |
| Some College | 32 (25.0) | 2968 (26.0) | 8 (21.6) | 2814 (25.4) | 9 (26.5) | 2569 (25.2) |
| Bachelor | 39 (30.5) | 2886 (25.3) | 14 (37.8) | 2856 (25.8) | 12 (35.3) | 2689 (26.3) |
| Post Graduate Degree | 45 (35.2) | 3862 (33.9) | 11 (29.7) | 3865 (34.9) | 11 (32.4) | 3583 (35.1) |
| **Marital Status** |  |  |  |  |  |  |
| Married | 87 (68.0) | 7673 (67.2) | 28 (75.7) | 7574 (68.0) | 24 (70.6) | 7036 (67.7) |
| Not Married | 41 (32.0) | 3648 (32.0) | 8 (21.6) | 3422 (31.4) | 10 (29.4) | 3115 (31.6) |
| **Family history - Psychosis** |  |  |  |  |  |  |
| No | 100 (78.7) | 9838 (87.2) | 32 (86.5) | 9522 (87.0) | 26 (76.5) | 8784 (87.0) |
| Yes | 24 (18.9) | 1147 (10.2) | 4 (10.8) | 1130 (10.3) | 8 (23.5) | 1046 (10.4) |
| **PQ-BC Distress score (mean(se))** | 8.34 (1.24) | 6.29 (0.10) | 3.86 (1.42) | 4.62 (0.09) | 4.09 (1.05) | 3.55 (0.08) |
| **CBCL Total Problems score (mean(se))** | 28.2 (2.38) | 17.9 (0.17) | 19.9 (3.07) | 17.5 (0.17) | 28.2 (4.01) | 16.4 (0.17) |

PQ-BC = Prodromal Questionnaire - Brief Child Version
CBCL = Child Behavior Checklist
SE = standard error

### 3.5. mTBI and PQ-BC Distress Scores

The negative binomial mixed-effects model did not find a strong association between mTBI and PQ-BC Distress scores (Table 4). Children with a possible mTBI had 1.17 times the rate (i.e., a 17% increased risk) of experiencing distress as a result of a PLE after adjustment for covariates (95% CI: 0.99, 1.40). There was a non-significant 14% increased risk of experiencing distress as a result of a PLE in children with mTBI (95% CI: 0.87, 1.49). In the models using imputed data (Table 5), possible mTBI was associated with a 21% increased risk (95% CI: 1.03, 1.43) of experiencing distress due to a PLE. The models further found a 22% increased risk of experiencing distress as a result of a PLE in those with a mTBI (95% CI: 0.95, 1.59).

**Table 4.** The Association Between Mild Traumatic Brain Injury and Scores on the PQ-BC and CBCL

|  | PQ-BC Distress |  | CBCL Total Problems |  |
| --- | --- | --- | --- | --- |
|  | IRR (95% CI) | p value | IRR (95% CI) | p value |
| **Traumatic Brain Injury** |  |  |  |  |
| No | Reference |  | Reference |  |
| Possible mild | 1.17 (0.99, 1.40) | 0.0711 | 1.07 (1.01, 1.13) | 0.0175 |
| Mild | 1.14 (0.87, 1.49) | 0.3520 | 1.15 (1.05, 1.26) | 0.0023 |

Note: Mixed-effects models adjusted for child age, gender, race/ethnicity, household educational attainment, household income, family history of psychosis, neighborhood safety, parent marital status, parental monitoring, family relationship.
IRR = Incidence Rate Ratio
CBCL = Child Behavior Checklist
PQ-BC = Prodromal Questionnaire – Brief Child Version

**Table 5.** Pooled parameter estimates for models using imputed data

|  | **PQ-BC Distress** |  | **CBCL Total Problems** |  |
| --- | --- | --- | --- | --- |
|  | IRR (95% CI) | p value | IRR (95% CI) | p value |
| **Traumatic Brain Injury** |  |  |  |  |
| No | Reference |  | Reference |  |
| Possible mild | 1.21 (1.03, 1.43) | 0.0218 | 1.08 (1.02, 1.14) | 0.0060 |
| Mild | 1.22 (0.95, 1.59) | 0.1239 | 1.19 (1.09, 1.30) | 0.0003 |

Note: Negative binomial mixed-effects models adjusted for child age, gender, race/ethnicity, household educational attainment, household income, family history of psychosis, neighborhood safety, parent marital status, parental monitoring, family relationship.
IRR = Incidence Rate Ratio
CBCL = Child Behavior Checklist
PQ-BC = Prodromal Questionnaire – Brief Child Version

### 3.6. mTBI and CBCL Total Problem Scores

The negative binomial mixed-effects model found a strong positive association between mTBI and CBCL Total Problems scores (Table 4). Children that experienced a mTBI had a 15% increased risk of an emotional or behavioral problem after adjustment for covariates (95% CI: 1.05, 1.26). Children with a possible mTBI experienced a 7% increased risk (95% CI: 1.01, 1.13) of an emotional or behavioral problem. The models using imputed data (Table 5) found an 8% (95% CI: 1.02, 1.14) and a 19% (95% CI: 1.09, 1.30) increased risk of an emotional or behavioral problem in children with a possible mTBI or mTBI, respectively.

### 3.7. Mediation Analyses

The mediation models included 7136 participants that attended the year-2 visit, did not have missing covariate data, and had MRI data that passed quality control. Missing MRI data were more likely from female participants, children belonging to a minority group, and children with missing household income (Supplementary Table S1). There was negligible evidence that sMRI metrics had a meaningful indirect effect between mTBI and behavioral outcomes (Table 6). Total cortical area mediated 1.27% of the association between mTBI and the PQ-BC Distress score (95% CI: -16.09, 18.63) and 1.29% of the association between mTBI and the CBCL Total Problems score (95% CI: -4.51, 7.08). Total cortical volume mediated 4.68% of the association between mBI and PQ-BC Distress score (95% CI: -33.08, 42.43) and 3.48% of the association between mTBI and CBCL Total Problems score (95% CI: -4.19, 11.15). Mean cortical thickness mediated 3.81% and 1.30% of the association between mTBI and PQ-BC Distress score or CBCL Total Problems score, respectively. Total subcortical gray volume mediated 2.68% of the association between mTBI and PQ-BC Distress score (95% CI: -29.09, 34.45). Whole brain volume mediated 3.15% and 2.37% of the association between mTBI and PQ-BC Distress score or CBCL Total Problems score, respectively. Indirect effects for all volumetric measures and PQ-BC Distress score ranged from -0.005 to 0.029. Proportion mediated ranged from -1.61% to 4.68%. Indirect effects for CBCL Total Problems score ranged from -0.002 to 0.020. Proportion mediated ranged from -0.43% to 3.48%.

**Table 6.**
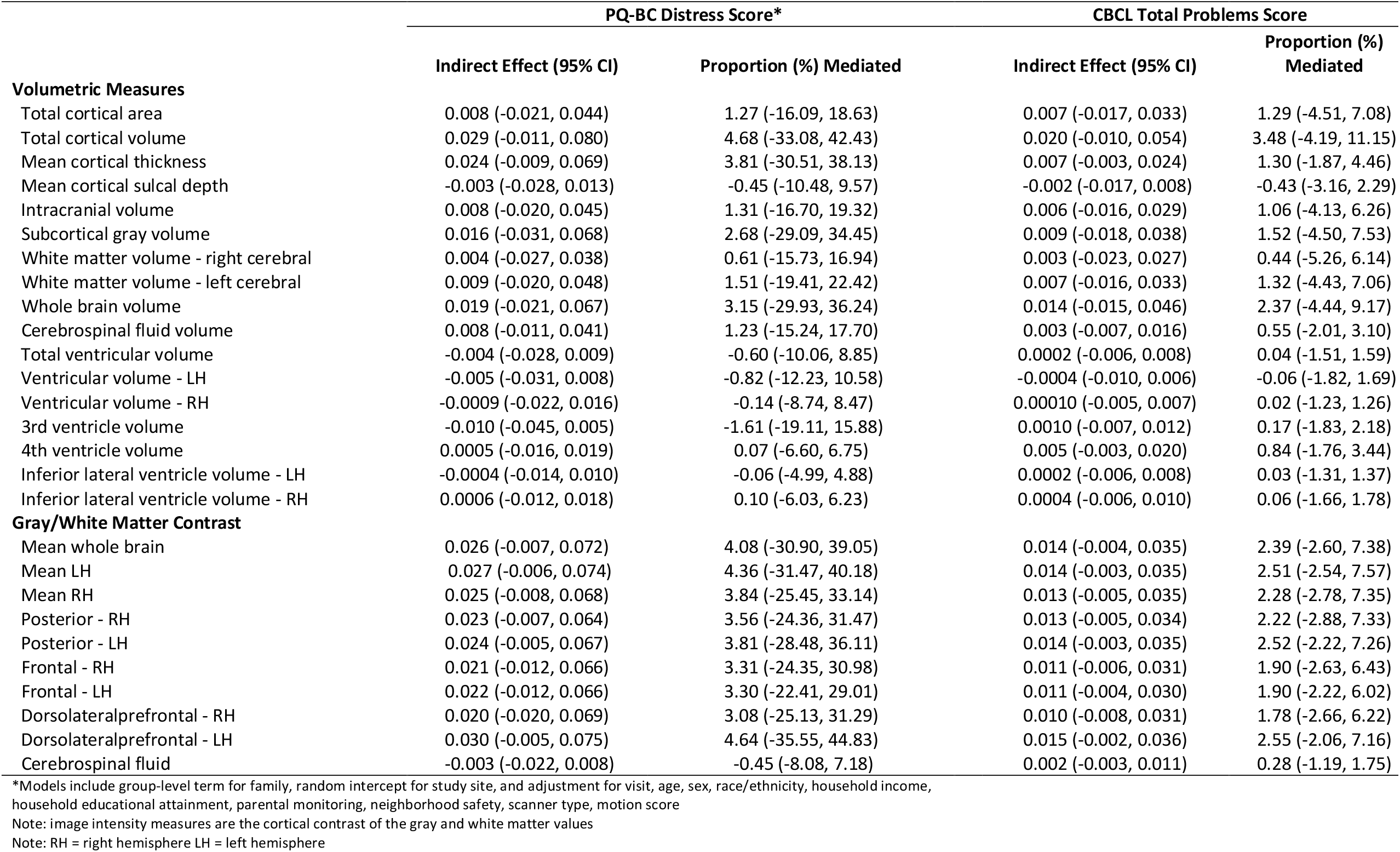
Results of Mediation Analyses for Mild TBI

Cortical contrast measures did not meaningfully mediate the association between mTBI and mental health outcomes (Table 6). The mean cortical GWC for the whole brain mediated 4.08% of the association between mTBI and PQ-BC Distress scores (95% CI: -30.09, 39.05) and 2.39% of the association between mTBI and CBCL Total Problems scores (95% CI: -2.60, 7.38). Indirect effects for all PQ-BC contrast measures ranged from -0.003 to 0.030. Indirect effects for all CBCL Total Problems contrast measures ranged from 0.002 to 0.015. Proportion mediated ranged from -0.45% to 4.64% for PQ-BC Distress scores and 0.28% to 2.55% for CBCL Total Problems scores.

#### 3.7.1. Associations Between mTBI and Volumetric Measures

There was evidence of an inverse association between mTBI and cerebrospinal fluid (CSF) volume when examining the path coefficients in the PQ-BC and CBCL mediation models (Table 7). In the PQ-BC and CBCL models, there was a meaningful decrease in CSF volume in children that had experienced a mTBI (β=-0.42, 95% CI: -0.83, -0.01). There was no evidence of a strong association for any other volumetric measure and mTBI.

**Table 7.** Path Effects for Mild TBI Mediation Analyses

|  | PQ-BC Distress Score** |  | CBCL Total Problems Score |  |
| --- | --- | --- | --- | --- |
|  | Association<br>between mild TBI<br>and ROI | Association between<br>ROI and PQ-BC | Association<br>between mild TBI<br>and ROI | Association between<br>ROI and CBCL |
| **Morphometric Measures** |  |  |  |  |
| Total cortical area | -0.09 (-0.41, 0.21) | -0.02 (-0.07, 0.03) | -0.10 (-0.40, 0.22) | -0.08 (-0.10, -0.05) |
| Total cortical volume | -0.22 (-0.52, 0.08) | -0.14 (-0.20, -0.08) | -0.21 (-0.53, 0.11) | -0.10 (-0.12, -0.07) |
| Mean cortical thickness | -0.23 (-0.55, 0.08) | -0.11 (-0.17, -0.05) | -0.23 (-0.54, 0.09) | -0.04 (-0.06, -0.01) |
| Mean cortical sulcal depth | 0.09 (-0.24, 0.41) | -0.05 (-0.11, 0.01) | 0.09 (-0.23, 0.43) | -0.03 (-0.06, -0.01) |
| Intracranial volume | -0.10 (-0.42, 0.21) | -0.09 (-0.16, -0.03) | -0.09 (-0.40, 0.22) | -0.07 (-0.10, -0.04) |
| Subcortical gray volume | -0.12 (-0.44, 0.22) | -0.15 (-0.21, -0.09) | -0.11 (-0.44, 0.22) | -0.08 (-0.11, -0.06) |
| White matter volume - right cerebral | -0.05 (-0.37, 0.29) | -0.09 (-0.15, -0.03) | -0.04 (-0.37, 0.31) | -0.07 (-0.10, -0.04) |
| White matter volume - left cerebral | -0.12 (-0.47, 0.22) | -0.09 (-0.15, -0.03) | -0.11 (-0.45, 0.22) | -0.07 (-0.10, -0.04) |
| Whole brain volume | -0.15 (-0.46, 0.16) | -0.13 (-0.20, -0.07) | -0.15 (-0.45, 0.16) | -0.10 (-0.12, -0.07) |
| Cerebrospinal fluid volume | -0.42 (-0.83, -0.01) | -0.02 (-0.07, 0.03) | -0.42 (-0.81, -0.04) | -0.01 (-0.03, 0.01) |
| Total ventricular volume | -0.17 (-0.56, 0.21) | 0.03 (-0.01, 0.08) | -0.17 (-0.58, 0.22) | -0.00 (-0.03, 0.02) |
| Ventricular volume - LH | -0.24 (-0.63, 0.15) | -0.00 (-0.23, 0.22) | -0.24 (-0.62, 0.14) | 0.00 (-0.02, 0.03) |
| Ventricular volume - RH | -0.05 (-0.44, 0.33) | 0.04 (-0.01, 0.09) | -0.05 (-0.43, 0.33) | -0.01 (-0.03, 0.02) |
| 3rd ventricle volume | -0.34 (-0.74, 0.05) | 0.04 (-0.01, 0.09) | -0.33 (-0.70, 0.04) | -0.00 (-0.03, 0.02) |
| 4th ventricle volume | -0.27 (-0.66, 0.12) | -0.00 (-0.05, 0.05) | -0.26 (-0.66, 0.13) | -0.02 (-0.05, 0.00) |
| Inferior lateral ventricle volume - LH | -0.05 (-0.44, 0.34) | 0.02 (-0.02, 0.06) | -0.05 (-0.44, 0.34) | -0.01 (-0.03, 0.01) |
| Inferior lateral ventricle volume - RH | 0.06 (-0.33, 0.45) | 0.03 (-0.02, 0.07) | 0.06 (-0.34, 0.45) | 0.01 (-0.01, 0.04) |
| **Gray/White Matter Contrast** |  |  |  |  |
| Mean whole brain | -0.17 (-0.40, 0.05) | -0.16 (-0.24, -0.08) | -0.18 (-0.40, 0.04) | -0.08 (-0.12, -0.05) |
| Mean LH | -0.19 (-0.42, 0.05) | -0.15 (-0.23, -0.07) | -0.19 (-0.42, 0.03) | -0.08 (-0.11, -0.05) |
| Mean RH | -0.16 (-0.37, 0.05) | -0.16 (-0.25, -0.08) | -0.16 (-0.12, 0.11) | -0.08 (-0.12, -0.05) |
| Posterior - RH | -0.15 (-0.36, 0.05) | -0.15 (-0.24, -0.07) | -0.16 (-0.37, 0.06) | -0.08 (-0.12, -0.05) |
| Posterior - LH | -0.18 (-0.41, 0.03) | -0.14 (-0.22, -0.06) | -0.18 (-0.40, 0.03) | -0.08 (-0.11, -0.05) |
| Frontal - RH | -0.14 (-0.36, 0.08) | -0.16 (-0.25, -0.08) | -0.14 (-0.36, 0.08) | -0.08 (-0.12, -0.05) |
| Frontal - LH | -0.15 (-0.37, 0.08) | -0.16 (-0.24, -0.08) | -0.15 (-0.34, 0.05) | -0.08 (-0.11, -0.05) |
| Dorsolateralprefrontal RH | -0.13 (-0.39, 0.12) | -0.17 (-0.24, -0.10) | -0.13 (-0.37, 0.10) | -0.08 (-0.11, -0.05) |
| Dorsolateralprefrontal LH | -0.20 (-0.43, 0.03) | -0.16 (-0.23, -0.08) | -0.20 (-0.43, 0.03) | -0.08 (-0.11, -0.05) |
| Cerebrospinal fluid | 0.13 (-0.08, 0.34) | -0.04 (-0.11, 0.04) | 0.13 (-0.09, 0.34) | 0.02 (-0.01, 0.05) |

\*Models include group-level term for family, random intercept for study site, and adjustment for visit, age, sex, race/ethnicity, household income, household educational attainment, parental monitoring, neighborhood safety, scanner type, motion score
Note: image intensity measures are the cortical contrast of the gray and white matter values
Note: RH = right hemisphere LH = left hemisphere

#### 3.7.2. Associations between Brain Measures and Mental Health Outcomes

Multiple brain measures were meaningfully associated with PQ-BC Distress scores and CBCL Total Problems scores in the mediation models (Table 7). In the PQ-BC models, the strongest association was found between subcortical gray volume (β=-0.15 95% CI: -0.21, -0.09) and PQ-BC Distress scores. In other words, a decrease in subcortical gray volume was associated with increased PQ-BC Distress scores. In the CBCL-Total Problems models, the strongest association was found in total cortical volume. Decreased total cortical volume was strongly associated with an increase in CBCL Total Problems score (β=-0.10, 95% CI: -0.12, -0.07).

## 4. Discussion

In the current study, we aimed to examine the association between mTBI, mental health outcomes, and structural brain metrics potentially mediating the relationship. We focused on structural brain metrics and image intensity measures (i.e., gray-white matter contrast) to examine if children with mTBI history had brain changes associated with poor mental health outcomes. First, we observed an increased risk of distress due to a PLE in children that had experienced a possible mTBI only in models using imputed data. We further found evidence of an increased risk for behavioral and emotional problems in children with a possible mTBI or mTBI. The associations remained significant after adjustment for confounding and multiple imputation for missing data. Second, we examined whether sMRI measures mediated the association between mTBI and mental health outcomes. We found that volumetric and image intensity measures did not meaningfully mediate the relation. Indirect effects and proportion mediated estimates were small for all examined measures. The indirect effect and proportion mediated estimates are consistent with findings from other ABCD studies examining earlier life exposures and poor mental health outcomes (53). Third, path effects found evidence of an inverse association between mTBI and CSF volume. Taken together, the study shows that experiencing a mTBI is significantly related to emotional and behavioral problems.

The finding that mTBI was significantly associated with poor mental health outcomes was not unexpected. Prior research has consistently reported increased risk of behavioral and emotional problems following TBI. A pediatric cohort study comparing children with mTBI and children with orthopedic injury reported significantly increased CBCL Total Problems scores in the former (OR = 3.00, 95% CI: 1.33, 6.77) (54). The risk was more pronounced in younger children (mean age = 10 years old) out to 1 year post injury. A systematic review of pediatric TBI and behavioral outcomes concluded that severity played a role in the persistence of behavioral problems (23). Likewise, we found an association between increasing severity of mTBI and CBCL Total Problems score. We found that children with possible mTBI and mTBI were at an 7% and 16% increased risk of developing behavioral and emotional problems when compared to children with no TBI, respectively. The association between mTBI and PLEs is a bit less clear. Our findings that a possible mTBI increased risk of distress due to PLEs more than an mTBI is contrary to intuition. Possible reasons for the unexpected results include misclassification of the exposure and bias due to missing data. Misclassification of exposure (i.e., TBI severity) can be due to information bias (e.g., failure to accurately recall a head injury) (55). We believe the misclassification was nondifferential with respect to the outcome (i.e., parent recall of the injury was not influenced by child reported distress due to PLEs) and likely attenuated mTBI results. Finally, results of the imputation models suggest that missing data biased our association downwards. The results are not surprising given the high PQ-BC Distress scores in children with missing household income (Table 2). The increased risk in the models with imputed data could be due to omission of a high-risk group with missingness. Future studies using ABCD data should examine differential patterns of missingness to address whether there is evidence of selection or missing-data bias.

We did not identify any evidence of meaningful mediation. Prior research using the ABCD dataset has found that brain ROIs accounted for significantly less variance than environmental and genetic factors (53). In our study, proportion mediated did not exceed 1-2% for any structural brain metrics except total cortical volume, mean cortical thickness, and subcortical gray volume. Proportion mediated for the GWC measures were generally greater in the PQ-BC Distress models, although the credible intervals were relatively wide (i.e., imprecise). The imprecision was likely influenced by the few numbers of children that endorsed the PQ-BC distress items at the year-2 visit. There are a few possibilities for the lack of meaningful mediation in our study. First, studies that have found significant structural MR changes after TBI often use very specific time ranges post injury to conduct the MRI scan, often occurring within 1 month of the injury (13, 56-58). Even within these studies, changing the time interval of MRI after injury resulted in different findings of GM and WM differences over time. Previous research has demonstrated that MR findings post TBI are dynamic rather than consistent, even moving in different directions depending on the time post-injury (57). The fact that this study does not have a way to control for time post-injury (i.e., the head injury reported at year-2 occurred at any time since the year-1 study visit) may be an explanation for the lack of meaningful mediation. Second, another possibility is the study definition for mTBI. The OTBI may have included a wider range of mild injuries. This is because the screener does not require a hospital or doctor visit to diagnose the injury, but solely relies on reported symptoms. In contrast, most studies conducted post-injury are using patients who report to a hospital or some medical professional shortly after the injury (13, 56-58). With this difference in inclusion criteria, the present study may be collecting a larger portion of mild injuries that have not been included in prior research and consequently have more mild or reduced brain changes.

Findings from the mediation analysis suggest that mTBI is associated with decreased CSF volume, although caution is warranted when interpreting the results. Abnormalities in CSF dynamics may be due to neurological injury (e.g., TBI, stroke) (59). Results of studies using samples with moderate and severe TBI find time-dependent alterations with respect to CSF volume. A prospective cohort study in an adult population with moderate or severe TBI reported a significant increase in CSF volume only in the post-acute period (i.e., more than 210 days after injury) (60). The findings emphasize the importance of incorporating time since injury. There are only a few studies examining CSF volume and mTBI. A longitudinal study with mild and moderate TBI patients did not detect any significant difference in the CSF volume of cases and controls (61). Another possibility for our unexpected finding (i.e., decreased CSF volume in children with an mTBI) is lesion-induced errors in the volumetric results. Prior research with pediatric populations and TBI have found biases in automated software packages (e.g., Freesurfer pipelines) that can be attributed to lesions (62). Methods that incorporate a focal pre-processing approach (e.g., enantiomorphic filling of the damaged area) can reduce errors and improve the detection of physiologically meaningful differences (62, 63).

The primary strength of the study is the longitudinal assessment of mental health outcomes in a large sample of children. The incorporation of three equally spaced time points allowed for an examination of mental health outcomes with respect to mTBI occurrence. The collection of a wide variety of instruments by the study limited the influence of confounding on our measures of association. Finally, we expect that study findings will be generalizable to a wide population due to the demographic makeup of the sample.

A limitation of our work is that we did not examine the longitudinal association of structural brain metrics with mTBI and mental health outcomes. First, the absence of MRI data at the year-1 visit posed a problem with interpretation of results (i.e., there was a gap in data collection that obscured neural trajectory). Second, the exclusion of baseline data from the mediation analysis was done due to concerns with recall bias with respect to injury severity and time since injury. Information bias is especially concerning considering the widely acknowledged underreporting of head injuries in minority populations (64). In our sample, we observed a disproportionately low number of mTBI in Black children (Table 3). Finally, the imaging findings potentially reflect a short-term consequence of mTBI rather than a lasting neurological impairment. The decision to restrict the mediation analysis to a single-time point (i.e., a cross-sectional analysis) limits claims of causal inference. The ongoing ABCD Study will further elucidate the longitudinal relation as more imaging data is collected.

## 5. Conclusion

In summary, these findings show that possible mTBI and mTBI is associated with an increased risk of poor behavioral and mental health outcomes in adolescents. Additional data is needed to identify meaningful brain mechanisms involved in the development of mental health challenges following mTBI.

## Data Availability

The ABCD data are openly available to qualified researchers for free. Access can be requested at https://nda.nih.gov/abcd/. Code for replication of the analyses conducted in this manuscript can be retrieved at https://github.com/Daniel-Adan-Lopez/ABCD_mTBI/blob/main/Rcode.

https://github.com/Daniel-Adan-Lopez/ABCD_mTBI/blob/main/Rcode

## Credit authorship contribution statement

**Daniel A. Lopez:** Conceptualization, Methodology, Formal analysis, Investigation, Writing – original draft, Writing – review & editing. **Zach P. Christensen:** Conceptualization, Methodology, Formal analysis, Investigation, Writing – review & editing. **John J. Foxe:** Conceptualization, Methodology, Investigation, Writing – review & editing. **Laura R. Ziemer:** Conceptualization, Investigation, Writing – original draft, Writing – review & editing. **Paige R. Nicklas:** Writing – original draft, Writing – review & editing. **Edward G. Freedman:** Conceptualization, Methodology, Investigation, Writing – original draft, Writing – review & editing.

## Abbreviations

ABCD: Adolescent Brain Cognitive Development
ROI: Region of interest
GWC: Gray/White Matter Contrast
CBCL: Child Behavior Checklist
PQ-BC: Prodromal Questionnaire – Brief Child Version
PLE: Psychotic-like experience
mTBI: mild Traumatic Brain Injury
IRR: Incidence Rate Ratio
GM: Gray Matter
WM: White Matter
sMRI: Structural Magnetic Resonance Imaging
CSF: Cerebrospinal Fluid
LOC: Loss of consciousness

---

**Table S1.** Characteristics of Participants Missing MRI at Year-2 Visit

|  | Missing MRI |  |
| --- | --- | --- |
|  | Yes (n=2543) | No (n=7845) |
| **Child gender** |  |  |
| Female | 1331 (52.3) | 3620 (46.1) |
| Male | 1212 (47.7) | 4225 (53.9) |
| **Race/ethnicity of child** |  |  |
| Asian | 67 (2.6) | 149 (1.9) |
| Black | 369 (14.5) | 1046 (13.3) |
| Hispanic | 561 (22.1) | 1519 (19.4) |
| White | 1276 (50.2) | 4315 (55.0) |
| Other | 270 (10.6) | 816 (10.4) |
| **Household Income (\$)** | | |
| ≤50k | 574 (22.6) | 1770 (22.6) |
| 50k to <100k | 540 (21.2) | 2033 (25.9) |
| 100k to <200k | 777 (30.6) | 2449 (31.2) |
| ≥200 | 379 (14.9) | 983 (12.5) |
| Missing | 273 (10.7) | 610 (7.8) |
| **Parent marital status** |  |  |
| Married | 1662 (66.3) | 5341 (68.2) |
| Not Married | 828 (33.0) | 2443 (31.2) |
| **Family history - psychosis** |  |  |
| No | 2200 (87.9) | 6725 (86.6) |
| Yes | 233 (9.3) | 835 (10.8) |
| **Neighborhood safety - youth** |  |  |
| Strongly Disagree | 50 (2.0) | 189 (2.4) |
| Disagree | 105 (4.1) | 378 (4.8) |
| Neutral | 394 (15.6) | 1449 (18.5) |
| Agree | 846 (33.4) | 2534 (32.3) |
| Strongly Agree | 1138 (44.9) | 3288 (41.9) |
| **PQ-BC Distress (mean (SD))** | 3.47 (7.76) | 3.59 (7.71) |
| **CBCL Total Problems (mean (SD))** | 15.57 (16.84) | 16.71 (17.37) |

